# Blood-based epigenome-wide analyses on the prevalence and incidence of nineteen common disease states

**DOI:** 10.1101/2023.01.10.23284387

**Authors:** Robert F. Hillary, Daniel L. McCartney, Elena Bernabeu, Danni A. Gadd, Yipeng Cheng, Aleksandra D. Chybowska, Hannah M. Smith, Lee Murphy, Nicola Wrobel, Archie Campbell, Rosie M. Walker, Caroline Hayward, Kathryn L. Evans, Andrew M. McIntosh, Riccardo E. Marioni

**Affiliations:** Centre for Genomic and Experimental Medicine, Institute of Genetics and Cancer, University of Edinburgh, Edinburgh, UK; Edinburgh Clinical Research Facility, University of Edinburgh, Edinburgh, UK; School of Psychology, University of Exeter, Exeter, UK; Medical Research Council Human Genetics Unit, Institute of Genetics and Cancer, University of Edinburgh, Edinburgh, UK; Division of Psychiatry, University of Edinburgh, Royal Edinburgh Hospital, Edinburgh, UK

**Author notes:** Correspondence: Riccardo E. Marioni.

**Keywords:** DNA methylation, health record linkage, epigenome-wide association studies, literature review, causal inference

## Abstract

**Background:** Blood DNA methylation can inform us about the biological mechanisms that underlie common disease states. Previous epigenome-wide analyses of common diseases often focus solely on the prevalence or incidence of individual conditions and rely on small sample sizes, which may limit power to discover disease-associated loci.

**Results:** We conduct blood-based epigenome-wide association studies on the prevalence of 14 common disease states in Generation Scotland (n_individuals_≤18,413, n_CpGs_=752,722). We also utilise health record linkage to perform epigenome-wide analyses on the incidence of 19 disease states. We present a structured literature review on existing epigenome-wide analyses for all 19 disease states to assess the degree of replication within the existing literature and the novelty of the present findings.

We identify 69 associations between CpGs and the prevalence of four disease states at baseline, of which 58 are novel. We also uncover 64 CpGs that associate with the incidence of two disease states (COPD and type 2 diabetes), of which 56 are novel. These associations were independent from common lifestyle risk factors. We highlight poor replication across the existing literature. Here, replication was defined by the reporting of at least one common gene in >2 studies examining the same disease state. Existing blood-based epigenome-wide analyses showed evidence of replication for only 4/19 disease states (with up-to-15% of unique genes replicated for lung cancer).

**Conclusions:** Our summary data and structured review of the literature provide an important platform to guide future studies that examine the role of blood DNA methylation in complex disease states.

## 1. Introduction

Epigenetic modifications to DNA represent an important mechanism by which the environment interacts with the genome (1). DNA methylation (DNAm) is one of the best-studied epigenetic mechanisms and involves the addition of methyl groups to DNA, typically in the context of cytosine-phosphate-guanine dinucleotides (CpG sites). In contrast to genetic sequence variation, these modifications are reversible and can modulate cell or tissue-specific patterns of gene expression (2). Genome-wide patterns of DNAm are most commonly assayed using microarray-based technologies such as the Illumina HumanMethylation 450K and HumanMethylationEPIC arrays. The arrays permit a cost-effective assessment of DNAm at a scale required for large-scale population health studies (3, 4).

Epigenome-wide association studies (EWAS) examine associations between the proportion of methylation at CpG sites and health outcomes of interest, such as chronic disease states (5). Primarily, EWAS have been conducted using whole-blood DNA methylation. Patterns of DNAm identified in blood do not necessarily mirror DNAm patterns in distal or disease-relevant tissues such as nervous tissue for Alzheimer’s disease (6, 7). However, blood sampling represents a minimally-invasive route for scalable biomarker measurement [10]. Blood-based EWAS have also implicated differential methylation at individual loci as candidate markers of disease risk. For example, *TXNIP* and *ABCG1* are important regulators of glucose and cholesterol metabolism, respectively. Hypomethylation within *TXNIP* (cg19693031) and *ABCG1* hypermethylation (cg06500161) have been associated with type 2 diabetes risk across individuals of multiple ancestries (8-11).

Existing EWAS on common disease can be categorised broadly into prevalence analyses (i.e. cross-sectional) and incidence analyses (i.e. longitudinal assessment of incident cases in unaffected individuals). EWAS often suffer from low sample sizes, which has limited the discovery of CpG sites that associate with disease states. Meta-analyses can increase power but may be vulnerable to between-study heterogeneities. To date, no study has performed large-scale EWAS on the prevalence and incidence of multiple disease states in a single population. There is also a lack of structured literature reviews on EWAS of common disease states to assess the level of agreement in loci discovery among existing studies.

Here, we utilise Generation Scotland: the Scottish Family Health Study (GS), a large cohort with DNA methylation data (n=18,413). First, we integrate blood DNAm and self-reported disease data from questionnaires answered at the study baseline to perform EWAS on 14 prevalent disease states (cross-sectional analyses). Second, we conduct EWAS on 19 incident disease states ascertained through electronic health record linkage over up-to-14 years of follow-up (longitudinal analyses). Third, we perform a structured literature review to identify blood-based EWAS findings on all 19 disease states considered in this study. We examine whether findings in this study replicate previous analyses and highlight the level of agreement within previously published studies. Fourth, we employ genetic colocalisation analyses to determine whether CpG methylation at loci identified in our EWAS causally associate with disease risk (see **Fig 1** for a summary of the study design).

**Fig 1.**
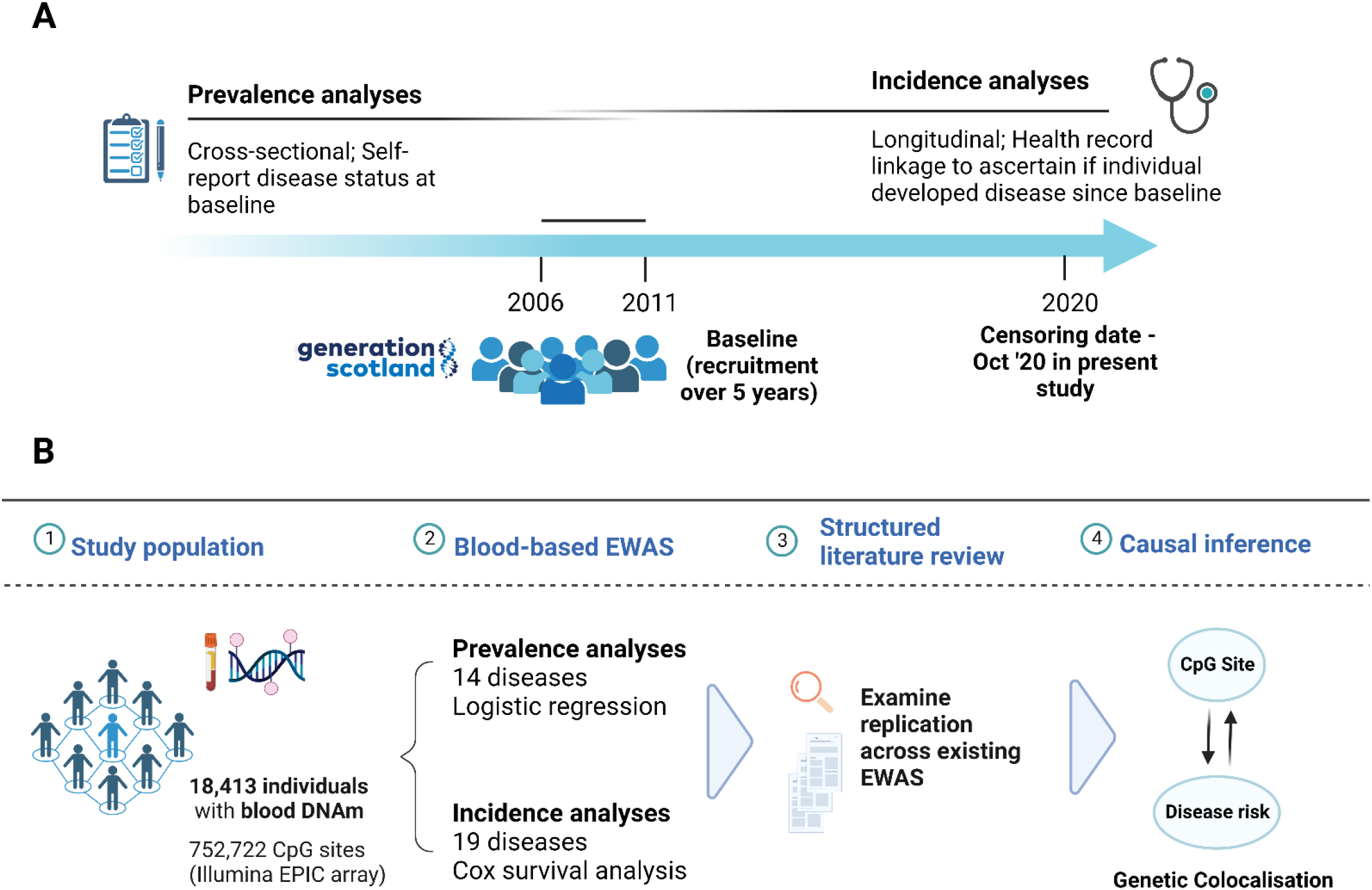
Study design for epigenome-wide analyses on prevalent and incident disease states in Generation Scotland. A). Recruitment for Generation Scotland took place between 2006 and 2011. *Prevalence analyses:* participants self-reported disease status and donated blood samples at the study baseline. *Incidence analyses:* linked healthcare data were used to determine if participants who were free from a particular condition at baseline went on to develop the condition over up-to-14 years of follow up. Controls were free of the disease at the baseline and during follow-up. B). Blood DNAm at baseline were available for 18,413 participants. EWAS tested for associations between blood CpG methylation and the prevalence of 14 disease states at baseline or the incidence (time-to-onset) of 19 disease states during follow-up. Significant findings were tested for replication in existing studies via a structured literature review. Replication within existing studies was also investigated. Causal inference analyses were employed to help dissect whether associations between DNAm and disease states reflected shared genetic architectures. Image created using Biorender.com. DNAm, DNA methylation; EWAS, epigenome-wide association studies.

## 2. Methods

### 2.1. Generation Scotland cohort

Generation Scotland, or GS, is a large family-structured cohort study that consists of 24,000 individuals from across Scotland. It encompasses 5,573 families with a median family size of 3 (interquartile range=2–5 members; excluding 1,400 singletons without any relatives in the study). Recruitment for GS took place between 2006 and 2011. Full details on the cohort and baseline data collection have been described previously (12, 13).

### 2.2. Preparation of DNA methylation data

Whole blood DNAm was measured for GS participants using the Illumina Infinium MethylationEPIC array. Dasen normalisation was performed across all individuals (14). Methylation M-values were corrected for age, sex and experimental batch (n=121 batches) prior to analyses. Methylation typing was performed in three distinct sets. Quality control steps are detailed in full in **Additional file 1**. Following quality control, there were 5,087, 4,450 and 8,876 individuals within Sets 1, 2 and 3, respectively. Set 1 contained related individuals. Set 2 consisted of individuals who were unrelated to each other and those in Set 1. Set 3 consisted of related individuals, and individuals related to those in Sets 1 and 2. In total, 752,722 probes and 18,413 individuals passed quality control criteria and were considered in analyses.

### 2.3. Preparation of disease phenotypes

Nineteen common disease states were considered across prevalence and incidence analyses: (1) Alzheimer’s dementia (AD), (2) breast cancer, (3) chronic kidney disease (CKD), (4) chronic neck and/or back pain, (5) chronic obstructive pulmonary disorder (COPD), (6) colorectal cancer, (7) COVID-19 severity (requiring hospitalisation), (8) inflammatory bowel disease (IBD), (9) ischemic heart disease, (10) liver cirrhosis, (11) long COVID, (12) lung cancer, (13) osteoarthritis, (14) ovarian cancer, (15) Parkinson’s disease, (16) prostate cancer, (17) rheumatoid arthritis, (18) stroke and (19) type 2 diabetes. Outcomes were selected if they were present among the ten leading causes of death in high-income countries, the ten leading causes of disease burden (disease adjusted life years; DALYs) in high-income countries or self-reported diseases at the baseline (15-17). Depression was not considered as it is included in an ongoing meta-analysis EWAS. Although asthma can occur at any age, it has a higher prevalence among children aged 0-17 years than in adults. It was therefore excluded from the present analyses that used an adult sample with a broad age profile (18).

Self-report data were used for 12 disease states in cross-sectional analyses of disease prevalence. Self-reported parental history of AD was used a proxy variable for AD. Analyses on self-reported parental history of AD were restricted to participants who were >45 years at baseline. This ensured that only participants whose parents were likely old enough at baseline to be at risk of AD were considered (i.e. >65 years). The CKD Epidemiology Collaboration, or CKD-EPI, equation was implemented to estimate glomerular filtration rate (eGFR) at baseline. Individuals with an eGFR <60 ml/min/1.73 m^2^ were deemed to have CKD (19). Therefore, 14 disease phenotypes were considered in prevalent analyses.

All 19 phenotypes were included in longitudinal analyses via linkage to electronic health records (with the exception of self-reported long COVID). The primary and secondary care codes used to define incident phenotypes are available in **Additional file 2**. Prevalent cases from the study baseline were excluded for these analyses as were those where record linkage provided evidence of a diagnosis prior to baseline. Therefore, incident cases included those diagnosed after baseline who had died and those who received a diagnosis and remained alive. Controls were censored if they were free of a diagnosis at the time of death or at the end of the follow-up period. Further information on the pre-processing of incident phenotypes, including COVID phenotypes, is available in **Additional file 1**.

### 2.4. Epigenome-wide association studies on prevalent disease

First, logistic regression models were used to adjust prevalent phenotypes for chronological age and sex, with the exception of breast cancer and prostate cancer, which were adjusted for age after restricting the cohort to females and males, respectively. Second, linear regression models were used for EWAS via the OSCA (OmicS-data-based Complex trait Analysis) software (20). Residuals from the logistic regression models were entered as the dependent variable and age-, sex- and batch-adjusted CpG M-values represented the independent variable. This strategy was employed to reduce computational burden. A Bonferroni-significance threshold was set at *p<*2.6×10^−9^ (=3.6×10^−8^/14 phenotypes) (21). Two models with different covariate strategies were employed, as described below:

1. **Basic model:** Phenotype and CpG M-values, corrected as described above, including five Houseman-estimated white blood cell (WBC) proportions as fixed effect covariates (22): Phenotype ∼ CpG M-values + methylation-predicted WBC proportions
2. **Fully-adjusted model:** additional adjustments for five common lifestyle factors (23) and 20 genetic principal components (PCs; to control for population structure): Phenotype ∼ CpG M-values + methylation-predicted WBC proportions + alcohol consumption (units/week) + body mass index (kg/m^2^) + deprivation index (Scottish Index of Multiple Deprivation) + education (an 11-category ordinal variable) + methylation-based smoking score (EpiSmokEr) + 20 genetic PCs

### 2.5. Epigenome-wide association studies on incident disease

First, Cox proportional-hazards models were used to adjust incident phenotypes for age at baseline and sex (17/19 phenotypes). Only age was included for breast, ovarian and prostate cancer. Time-to-onset for the disease, or censoring, was the survival outcome in Cox proportional-hazards models. Only individuals with an age at event or censoring ≥65 years were considered for AD. Logistic regression models were used to adjust two remaining COVID phenotypes prior to EWAS analyses. Cox models were not employed for COVID phenotypes owing to the limited differences in time-to-event data between individuals with positive COVID diagnoses. Whereas DNAm was corrected for age at baseline (as well as sex and batch), COVID phenotypes were adjusted for sex and age at COVID testing or diagnosis. Here, age at COVID testing or diagnosis was considered given the variation in time elapsed between baseline visits (between 2006 and 2011) and the onset of the COVID pandemic. Second, martingale residuals or logistic regression residuals were extracted and included as dependent variables in OSCA. A Bonferroni-corrected significance threshold was set at *p*<1.9×10^−9^ (=3.6×10^−8^/19 phenotypes). Basic and fully-adjusted models were employed, as described in the previous section. Methods for sensitivity EWAS analyses are detailed under **Additional file 1**.

### 2.6. Structured literature review on blood-based EWAS of common disease

MEDLINE, Embase (Ovid interface, 1980 onwards), Web of Science (core collection, Thomson Reuters), and preprint servers were searched to identify relevant articles indexed as of August 31, 2022. Search terms are outlined under **Additional file 1**. The search strategy returned approximately unique 2,000 articles, of which 53 passed inclusion criteria. Inclusion criteria were as follows: (i) original research article, (ii) EWAS performed with blood DNAm, (iii) there were at least 20 individuals in each comparison group (i.e. cases and controls) and (iv) the study examined at least one of the 19 common disease states outlined in our study.

### 2.7. Colocalisation analyses

Colocalisation analyses required GWAS summary statistics for CpG sites (i.e. methylation Quantitative Trait Loci – mQTLs, trait 1) and for respective disease states (trait 2 (24-29)). The GoDMC mQTL resource represents the largest mQTL study to date in terms of sample size but only focused on 450k array sites (30). Therefore, the GoDMC resource was utilised for sites in our study that are common to the EPIC and 450k arrays; however, mQTL analyses were also conducted in GS in order to generate mQTL summary statistics for sites present on the EPIC array only (**Additional file 1**). The coloc.abf function in R package *coloc* was used to test for colocalisation and default parameters were applied (version 5.1.0) (31). SNPs ±1 Mb surrounding each CpG site were extracted from the mQTL and respective disease GWAS summary statistics. The method tests for five mutually exclusive hypotheses, H0: there are no causal variants for either trait in the tested region; H1 and H2: causal variant for trait 1 and trait 2 only, respectively; H3: distinct causal variants for both traits and H4: the traits share a causal variant. Posterior probabilities ≥95% for H4 provided strong evidence in favour of colocalisation.

## 3. Results

### 3.1. Demographics in Generation Scotland

The mean age of the sample was 47.5 years (n=18,413, standard deviation (SD)=14.9) and the sample was 58.8% female. Summary data for demographic variables and disease counts are displayed in **Additional file 3: Tables S1-S3**. Associations between covariates and disease states are displayed in **Additional file 3: Tables S4 and S5** for prevalent and incident disease states, respectively (also available in **Additional file 4: Fig S1 and S2**).

### 3.2. Epigenome-wide analyses of prevalent disease

We first tested for cross-sectional associations between blood CpG methylation and fourteen disease states at the study baseline. There were 1,340 significant associations involving ten diseases in a basic model that adjusted for age, sex and estimated blood cell proportions (*p*<2.6×10^−9^, **Fig 2A, Additional file 3: Table S6**). Over 90% of these associations (n=1,246) were attributed to type 2 diabetes (n=703 associations, 52.2%), COPD (n=301, 22.5%) and chronic pain (n=242, 18.1%). Look-up analyses in the EWAS Catalog showed that 617/1,340 associations involve CpGs that were previously associated with common disease risk factors including body mass index, smoking and alcohol consumption (32).

**Fig 2.**
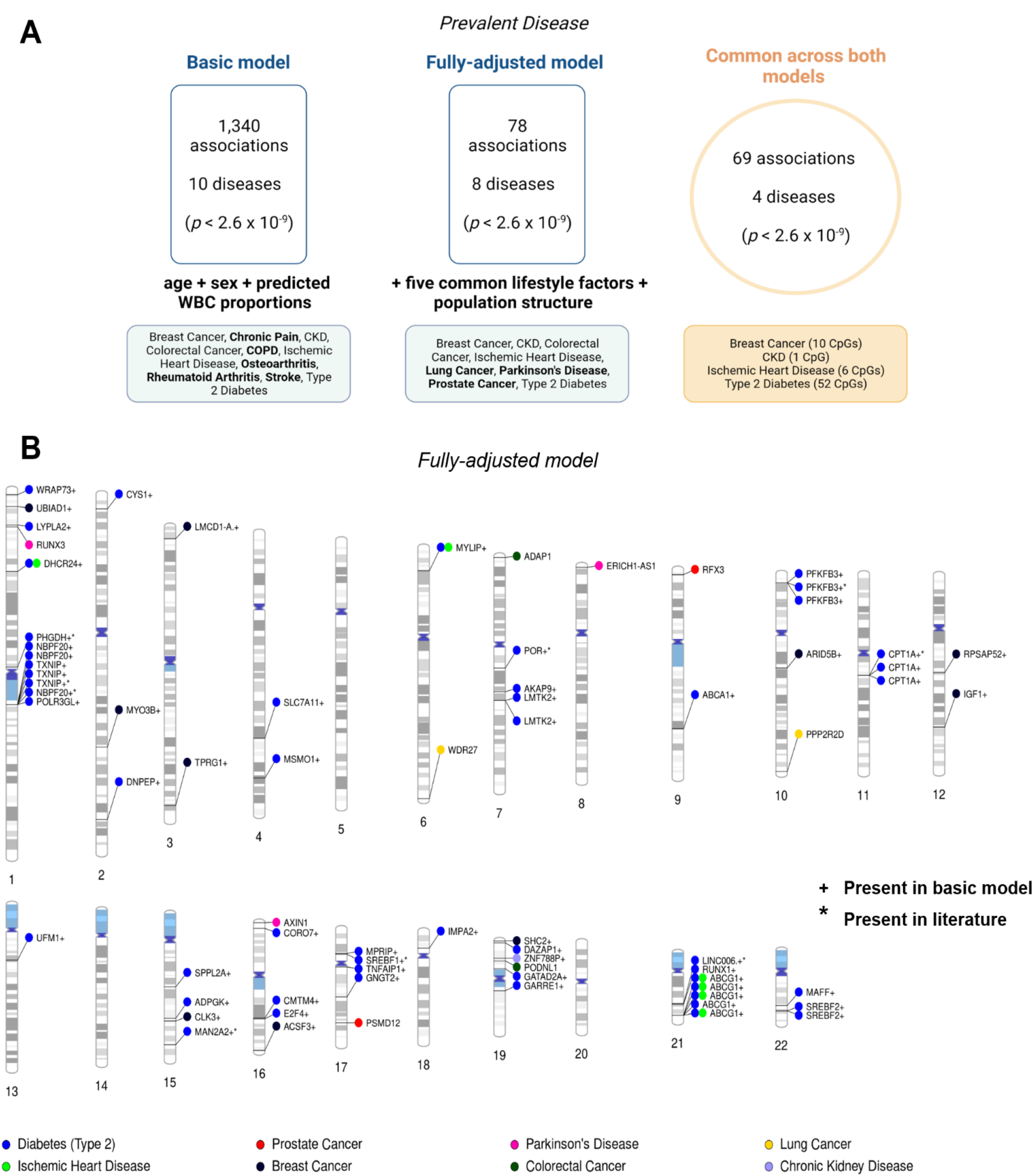
Epigenome-wide association studies on 14 prevalent disease states in Generation Scotland. A). Diseases with CpG associations in only the basic model or the fully-adjusted model are shown in bold. Colorectal cancer was present in both the basic and fully-adjusted model but no CpGs were common to both models for this condition. B). Ideogram showing 78 Bonferroni-corrected significant loci from the fully-adjusted model, which associated with 8 unique disease states. Loci are denoted with ‘*’ if they were replicated in the literature and with ‘+’ if they were also present in the basic model. Gene names that are greater than 10 characters in length were truncated for clarity. Full information is available in **Additional file 3: Table S7**. Image created using Biorender.com. CKD, chronic kidney disease; COPD, chronic obstructive pulmonary disease; WBC, white blood cells.

Next, we conducted a fully-adjusted model that further accounted for five common lifestyle risk factors and population structure, which returned 78 associations across 8 disease states (*p*<2.6×10^−9^, **Fig 2B, Additional file 3: Table S7**). Sixty-nine associations from the basic model were also present in the fully-adjusted analysis. The 69 associations were spread across four disease states: CKD (n=1); ischemic heart disease (n=6); breast cancer (n=10); and type 2 diabetes (n=52). The findings included associations between self-reported history of breast cancer and hypomethylation within cg06072257 and cg06123699, which are located near *UBIAD1 and TPRG1* on chromosomes 1 and 3, respectively (*p=*6.5×10^−103^ and *p=*2.4×10^−101^, respectively). The site cg17944885 located between *ZNF788* and *ZNF20* on chromosome 19 associated with prevalent CKD (*p*=1.7×10^−12^). Furthermore, CpGs annotated to *ABCG1, DHCR24 and MYLIP* were common to ischemic heart disease and type 2 diabetes (**Fig 2B**).

Genetic colocalisation analyses provided weak evidence for a shared causal variant underlying methylation at cg00857282 (*MYLIP*) and risk of ischemic heart disease (PP=63%, **Additional file 3: Table S8**). There was also moderate evidence for distinct causal variants underlying ten of the 69 prevalent associations (PP>75%).

### 3.3. Epigenome-wide analyses on incident disease

Using health record linkage, we tested whether CpGs measured at baseline associated with the future onset of 19 disease states. We observed 14,047 associations between baseline CpG methylation and the incidence of 11 disease states in the basic model (*p*<1.9×10^−9^, **Fig 3A, Additional file 3: Table S9**). Of these, 11,305 (80.4%) and 2,657 (18.9%) were attributed to COPD and type 2 diabetes, respectively. Well-established smoking-associated probes (e.g. cg14391737 within *PRSS23* and cg05575921 within *AHRR*) associated with the incidence of COPD, lung cancer, ischemic heart disease, stroke, pain and/or CKD.

**Fig 3.**
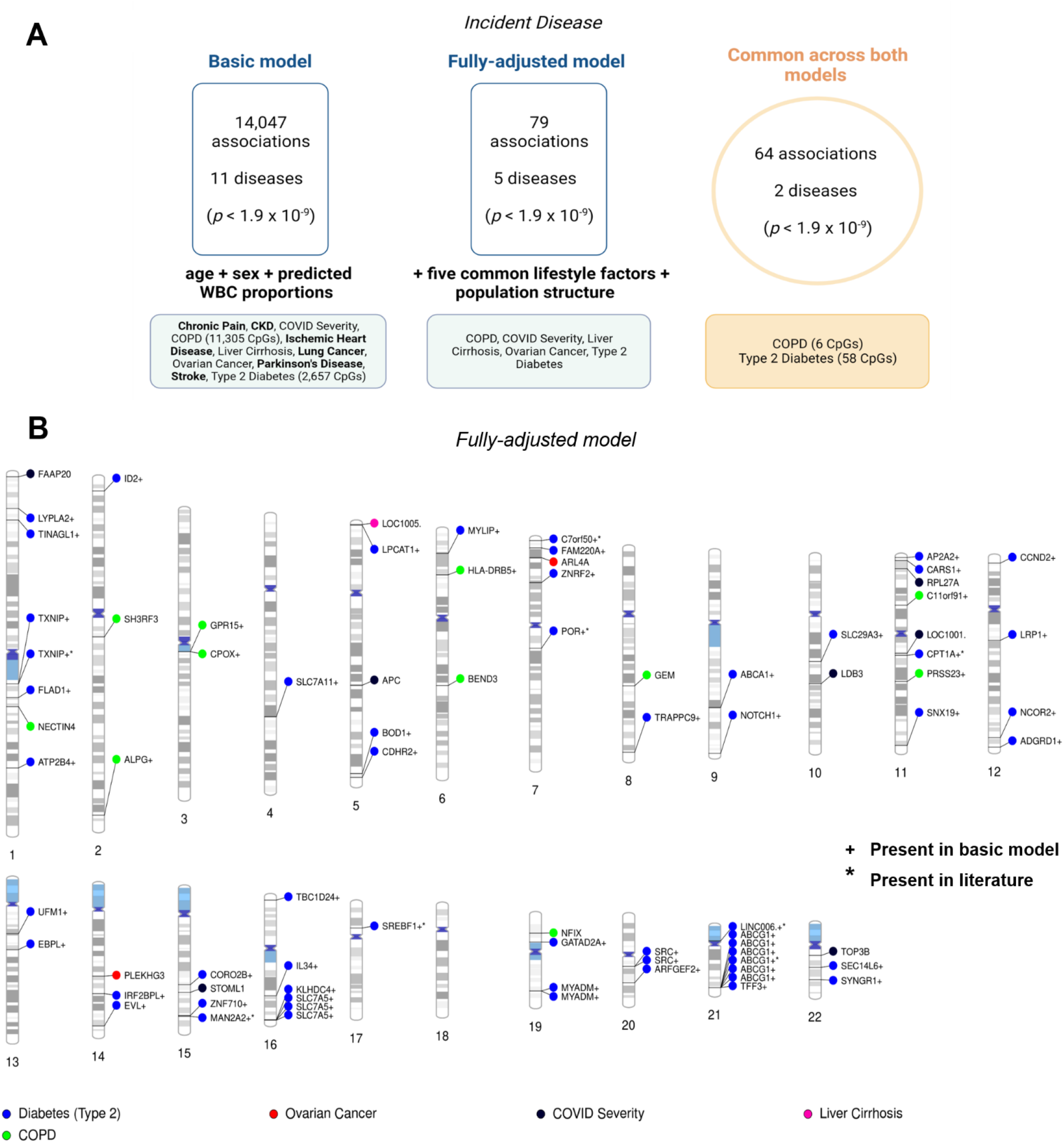
Epigenome-wide association studies on 19 incident disease states in Generation Scotland. Diseases that were identified in only the basic model or only the fully-adjusted model are shown in bold. COVID severity, liver cirrhosis and ovarian cancer were present in both a basic and fully-adjusted model but there were no overlapping CpGs for these disease states in both models. B). Ideogram showing 79 Bonferroni-corrected significant loci from the fully-adjusted model, which associated with 5 unique disease states. Loci are denoted with ‘*’ if they were replicated in the literature and with ‘+’ if they were also present in the basic model. Gene names that are greater than 10 characters in length were truncated for clarity. Full information is available in **Additional file 3: Table S10**. Image created using Biorender.com. COPD, chronic obstructive pulmonary disease; WBC, white blood cells.

There were 79 unique associations in the fully-adjusted model, which were spread across five disease states (**Fig 3B, Additional file 3: Table S10**). However, only 64 associations for COPD (n=6) and type 2 diabetes (n=58) were present across both basic and fully-adjusted models. Genes annotated to CpGs that associated with COPD included *ALPG, C11orf91, CPOX, GPR15, HLA-DRB5* and *PRSS23*. Genes annotated to CpGs that were associated with type 2 diabetes included *ABCA1, ABCG1, CPT1A, SREBF1, SLC7A11, SLC7A5, and TXNIP* among others (see **Additional file 3: Table S10** for full details). Only type 2 diabetes had CpGs common to cross-sectional and longitudinal analyses and reflected 17 CpGs annotated to 11 unique genes.

There was only moderate evidence for distinct causal variants underlying 11/64 incident associations (PP>75%). No associations showed strong evidence of colocalisation (**Additional file 3: Table S11**).

### 3.4. Associations between CpG methylation and disease states are robust in sensitivity analyses

Mixed-effects models that included a kinship matrix were used to account for relatedness as sensitivity analyses. Effect sizes correlated >0.99 with associations from the standard EWAS, which included related individuals (**Additional file 3: Tables S12 and S13, Additional file 4: Fig S3**). Further, fourteen of the 64 incident associations failed the proportional hazard assumption for the CpG variable (p<0.05 between Schoenfeld residuals and time, **Additional file 3: Table S14**). However, there were negligible differences between CpG effect sizes between follow-up periods that satisfied the assumption versus those that did not (**Additional file 3: Table S15**).

Fully-adjusted models were repeated using logistic regression (prevalent disease) or Cox models (incident disease) with age and sex included as fixed-effect covariates. This differs from the main analytical strategy that used linear regression models with adjusted phenotype and methylation variables, and allowed us to return effect sizes on an interpretable scale. **Fig 4** shows odds ratios and hazard ratios associated with a per-1 SD increase in adjusted CpG methylation M-values for all 69 and 64 prevalent and incident disease associations (**Additional file 3: Table S16**).

**Fig 4.**
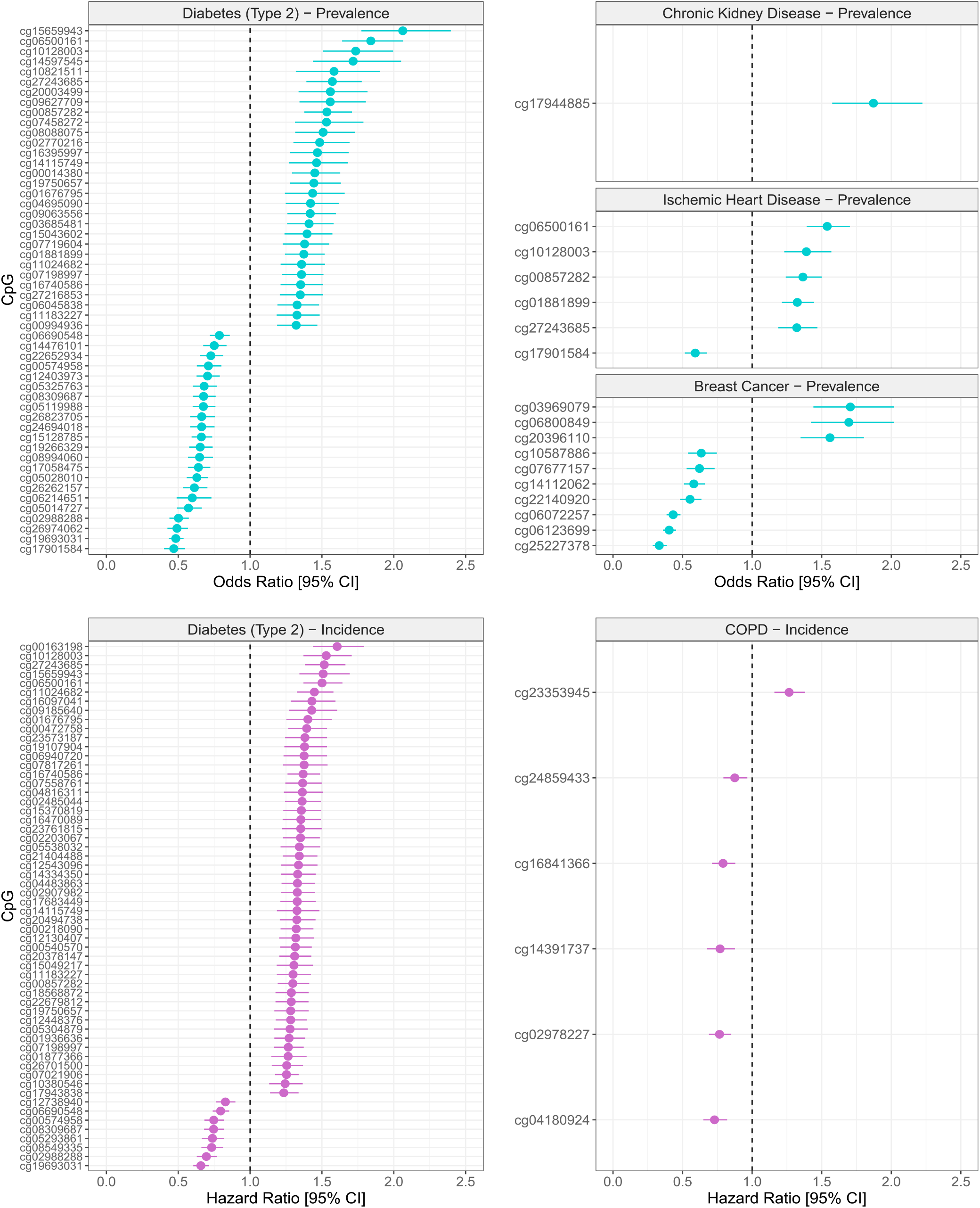
Blood CpGs associated with prevalent or incident disease states showing effect sizes on interpretable scale. Effect sizes were re-estimated using logistic regression (prevalent disease, blue points) or Cox proportional-hazards models (incident disease, violet points) to return more interpretable effect sizes. Effect sizes represent a per-1 standard-deviation increase in age-, sex- and experimental batch-adjusted CpG methylation M-values (or age- and batch-adjusted for breast cancer). CpGs shown were significant in both basic and fully-adjusted models. Odds ratios and hazard ratios are detailed in **Additional file 3: Table S16**.

### 3.5. Structured literature review on existing epigenome-wide analyses of common diseases

We performed a structured review of the literature to identify blood-based EWAS on the 19 disease states considered in our study (n=53 studies, **Fig 5A**). Fourteen disease states had at least one EWAS reported in the literature. The number of studies ranged from one (for long COVID) to seven (for type 2 diabetes) (**Additional file 3: Tables S17 and S18**).

**Fig 5.**
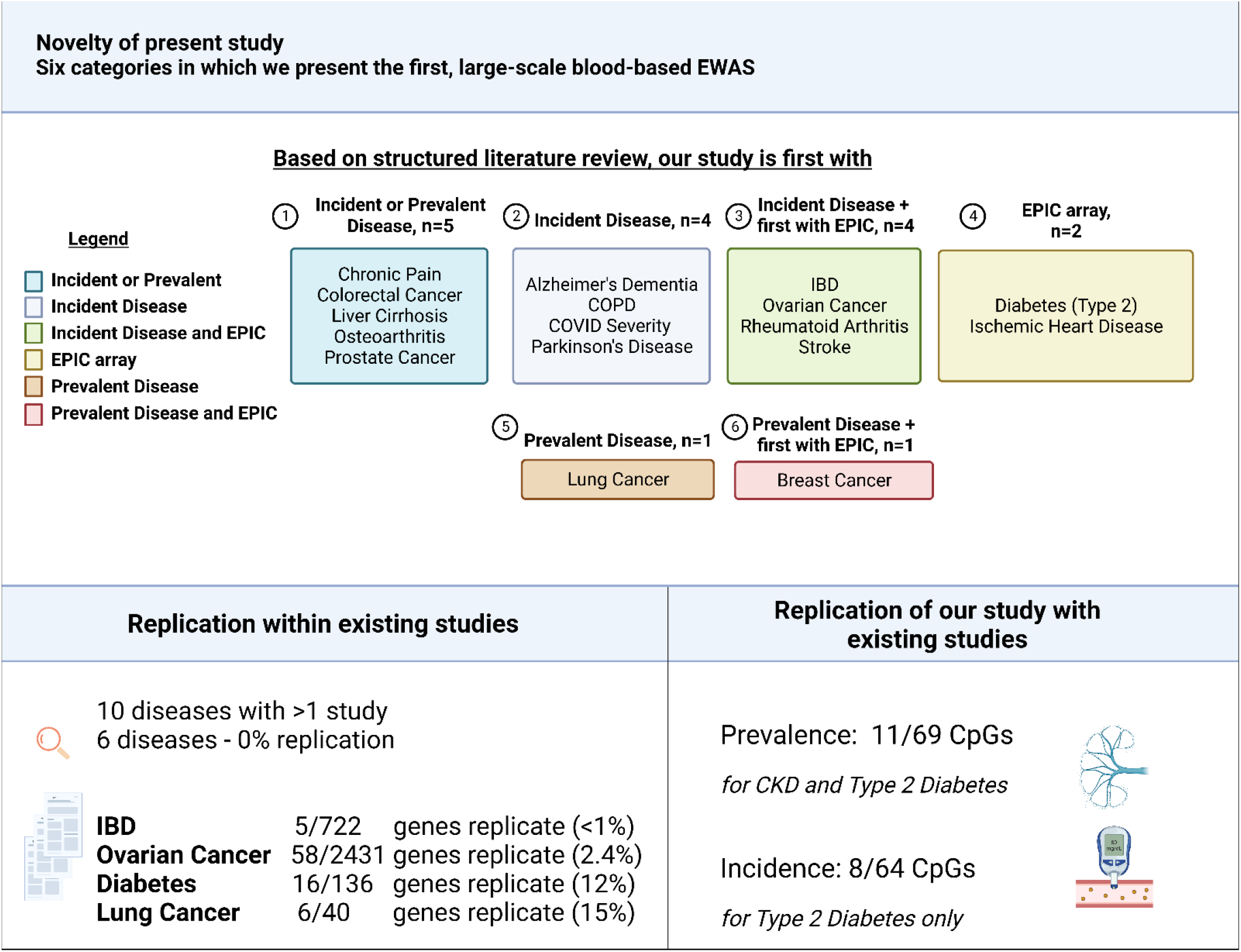
Summary of novelty of present study and poor replication within existing EWAS on common disease states. In some cases, there is an existing blood-based EWAS that examined the prevalence of the disease but no EWAS exists that analysed the incidence of the disease, and vice versa. In some cases, we present the first large-scale, blood-based EWAS of the disease. Further, few studies have performed EWAS with the recent Illumina EPIC array. Owing to these criteria, we define six possible categories in which we present the first EWAS on disease. *Interpretation:* For example, we present the first incident analyses on 4 diseases (category 2) but all 4 have at least one existing study examining prevalent disease and with the EPIC array. For another 4 diseases (category 3), we present the first EWAS on incident disease and the first with the EPIC array as any existing studies examining the prevalence of this disease have used earlier arrays only. We also highlight in a structured literature review the poor replication that exists between blood-based EWAS on disease, which extends to limited replication for our study. Image created using Biorender.com. CKD, chronic kidney disease; IBD, inflammatory bowel disease.

There were 10 disease states that were available for testing (i.e. had >1 study with complete summary statistic data). The number of unique CpGs that were reported as significant by the study authors ranged from 7 (for COPD) to 2,746 (for ovarian cancer). Four of the ten disease states had evidence of replication across existing studies with respect to the genes identified by EWAS. They were IBD (0.69% of genes replicated), ovarian cancer (2.4%), type 2 diabetes (12.0%) and lung cancer (15.0%) (**Fig 5B**). Whereas, CKD had no common genes identified across existing studies, 6/115 unique intergenic CpGs were replicated across studies (**Additional file 3: Table S19**).

Only 11/69 prevalent associations in this study (including 1 for CKD and 10 for type 2 diabetes) and 8/64 incident associations (for type 2 diabetes only) were replicated in in the literature (at *p*<2×10^−5^, which represented the least conservative threshold across studies for these traits). The replicated associations for type 2 diabetes implicated genes including *ABCG1, CPT1A, SREBF1* and *TXNIP*.

## Discussion

Using one of the world’s largest methylation datasets, we perform a series of EWAS on the prevalence and incidence of a broad range of conditions. We undertook a large-scale, comprehensive review of the literature and highlight the poor agreement that exists across previous epigenome-wide analyses that examine the same condition. By comparing these data with our own findings, we uncover 58 novel associations with the prevalence of three self-reported disease states at the study baseline (breast cancer, ischemic heart disease and type 2 diabetes). We also identify 56 novel associations between CpGs and the time-to-onset of two disease states (COPD and type 2 diabetes). These associations were independent of common lifestyle risk factors. However, we also observe a vast number of additional associations whereby CpGs index or track associations between lifestyle factors and common disease states, further highlighting the appropriateness of DNAm as a biomarker of lifestyle behaviours.

The novel associations observed in this study could strengthen evidence for candidate molecular pathways underlying peripheral disease states. For instance, self-reported history of breast cancer associated with differential methylation at cg06072257 (*UBIAD1*) and cg06123699 (*TPRG1*). UBIAD1 (UbiA Prenyltransferase Domain Containing 1) is a biosynthetic enzyme that converts vitamin K1 (phylloquinone) to menaquinone, which is the most abundant form of vitamin K2 in human tissue (33). Low expression of UBIAD1 in human breast tumours correlates with reduced survival (34), and also associates with risk for bladder cancer (35). *TPRG1* encodes for Tumour protein P63 Regulated 1 and its expression is associated with estrogen receptor-positive and triple-negative breast cancers (36, 37). *IGF1* and *SHC2* hypomethylation correlated with breast cancer prevalence in the present study. Whereas it is unclear how *SHC2* (chromosome 19) relates to cancer risk, *SHC1* (chromosome 1) and *IGF1* play central and causal roles in the pathogenesis of breast cancer (38-40). Furthermore, in relation to COPD, cg23353945 (*C11orf91*) correlated with incidence of the disease and has been associated in *trans* with CCL21 protein levels (41). Serum CCL21 levels are elevated in COPD patients and may contribute to the development of lung cancer (42, 43). This may suggest that a C11orf91-CCL21 axis contributes to risk of pulmonary disease independently from lifestyle risk factors. However, these findings warrant further investigation in mechanistic *in vitro* and *in vivo* studies.

The poor replication across existing studies reflects a number of possible factors. These include the use of different significance thresholds, arrays with different CpG content (e.g. 450k vs. EPIC arrays), different study designs (e.g. community-based designs with no enrichment for a particular disease vs. targeted case/control designs), variation in phenotype definitions for health record linkage analyses and the use of disparate covariate strategies. Some studies also did not make full summary statistics available. Nevertheless, our review is critical and timely given that the scale of EWAS continues to rise in tandem with enhancements in array technologies, population biobank sizes and health record phenotyping algorithms. We recommend that studies examining the same condition should reach consensus on covariate strategies in consortium efforts or report clearly the output of nested models such as models with and without adjustments for lifestyle risk factors.

Our study has a number of limitations. First, we did not adjust for medication data, which may confound associations between peripheral methylation and disease. Second, we did not consider disease subtypes as this may have reduced power to detect associations. Third, our findings in blood might not reflect important changes in distal, disease-relevant tissues. Fourth, our analyses consisted of individuals with European ancestry and might not be generalisable to individuals of other ancestries.

## Conclusions

Our epigenome-wide analyses uncovered over 100 novel associations between blood CpGs and common disease states that act independently of major confounding risk factors. Our summary data and synthesis of the literature provide a timely foundation that will expedite discoveries into the role of blood DNA methylation in common disease states.

## Supporting information

Additional file 2 - Disease Code Lists

## Data Availability

According to the terms of consent for Generation Scotland participants, access to data must be reviewed by the Generation Scotland Access Committee. Applications should be made to.
All code associated with this manuscript is available open access at the following GitHub repository: https://github.com/marioni-group/EWAS-of-common-disease-Generation-Scotland
EWAS summary statistics will be submitted to the EWAS Catalog and Edinburgh DataShare upon publication. 

## Availability of data and material

According to the terms of consent for Generation Scotland participants, access to data must be reviewed by the Generation Scotland Access Committee. Applications should be made to.

All code associated with this manuscript is available open access at the following GitHub repository: https://github.com/marioni-group/EWAS-of-common-disease-Generation-Scotland

EWAS summary statistics will be submitted to the EWAS Catalog and Edinburgh DataShare upon publication.

## Ethics approval and consent to participate

All components of GS received ethical approval from the NHS Tayside Committee on Medical Research Ethics (REC Reference Number: 05/S1401/89). GS has also been granted Research Tissue Bank status by the East of Scotland Research Ethics Service (REC Reference Number: 20-ES-0021), providing generic ethical approval for a wide range of uses within medical research.

## Supporting information captions

- **Additional file 1** – Supplementary Methods.
- **Additional file 2** – Disease code lists.
- **Additional file 3** – Supplementary Tables. **Table S1**. Summary data for demographic variables and covariates. **Table S2**. Counts for prevalent disease states. **Table S3**. Counts for incident disease states. **Table S4**. Associations between covariates and prevalent disease states. **Table S5**. Associations between covariates and incident disease states. **Table S6**. EWAS on prevalent disease - basic model. **Table S7**. EWAS on prevalent disease - fully-adjusted model. **Table S8**. Colocalisation analyses between blood CpGs and disease – prevalence. **Table S9**. EWAS on incident disease - basic model. **Table S10**. EWAS on incident disease - fully-adjusted model. **Table S11**. Colocalisation analyses between blood CpGs and disease – incidence. **Table S12**. Sensitivity - relatedness analyses - prevalent disease states. **Table S13**. Sensitivity - relatedness analyses - incident disease states. **Table S14**. Sensitivity - Cox proportional hazard assumptions. **Table S15**. Sensitivity - Hazard ratios per year for those failing tests in Table S14. **Table S16**. Odds ratios and hazard ratios for prevalent and incident disease associations. **Table S17**. Studies identified in structured literature review. **Table S18**. Contribution of present study to novelty. **Table S19**. Replication within the existing EWAS on disease.
- **Additional file 4** – Supplementary Figures. **Fig S1**. Associations between covariates and prevalent disease states in univariate and multivariate logistic regression models. **Fig S2**. Associations between covariates and incident disease states in univariate and multivariate Cox proportional-hazards models. **Fig S3**. Correlation between effect sizes from linear regression EWAS and sensitivity linear mixed effects analyses that further accounted for relatedness.

## References

1. Beck S, Rakyan VK. The methylome: approaches for global DNA methylation profiling. Trends in genetics : TIG. 2008;24(5):231–7.

2. Jaenisch R, Bird A. Epigenetic regulation of gene expression: how the genome integrates intrinsic and environmental signals. Nature genetics. 2003;33(3):245–54.

3. Bibikova M, Barnes B, Tsan C, Ho V, Klotzle B, Le JM, et al. High density DNA methylation array with single CpG site resolution. Genomics. 2011;98(4):288–95.

4. Pidsley R, Zotenko E, Peters TJ, Lawrence MG, Risbridger GP, Molloy P, et al. Critical evaluation of the Illumina MethylationEPIC BeadChip microarray for whole-genome DNA methylation profiling. Genome biology. 2016;17(1):1–17.

5. Flanagan JM. Epigenome-wide association studies (EWAS): past, present, and future. Methods in molecular biology (Clifton, NJ). 2015;1238:51–63.

6. Hannon E, Lunnon K, Schalkwyk L, Mill J. Interindividual methylomic variation across blood, cortex, and cerebellum: implications for epigenetic studies of neurological and neuropsychiatric phenotypes. Epigenetics. 2015;10(11):1024–32.

7. Gadd DA, Stevenson AJ, Hillary RF, McCartney DL, Wrobel N, McCafferty S, et al. Epigenetic predictors of lifestyle traits applied to the blood and brain. Brain Communications. 2021;3(2).

8. Meeks KAC, Henneman P, Venema A, Addo J, Bahendeka S, Burr T, et al. Epigenome-wide association study in whole blood on type 2 diabetes among sub-Saharan African individuals: findings from the RODAM study. International journal of epidemiology. 2019;48(1):58–70.

9. Juvinao-Quintero DL, Marioni RE, Ochoa-Rosales C, Russ TC, Deary IJ, van Meurs JBJ, et al. DNA methylation of blood cells is associated with prevalent type 2 diabetes in a meta-analysis of four European cohorts. Clinical Epigenetics. 2021;13(1):40.

10. Chambers JC, Loh M, Lehne B, Drong A, Kriebel J, Motta V, et al. Epigenome-wide association of DNA methylation markers in peripheral blood from Indian Asians and Europeans with incident type 2 diabetes: a nested case-control study. The lancet Diabetes & endocrinology. 2015;3(7):526–34.

11. Fraszczyk E, Spijkerman AMW, Zhang Y, Brandmaier S, Day FR, Zhou L, et al. Epigenome-wide association study of incident type 2 diabetes: a meta-analysis of five prospective European cohorts. Diabetologia. 2022;65(5):763–76.

12. Smith BH, Campbell H, Blackwood D, Connell J, Connor M, Deary IJ, et al. Generation Scotland: the Scottish Family Health Study; a new resource for researching genes and heritability. BMC medical genetics. 2006;7:74.

13. Smith BH, Campbell A, Linksted P, Fitzpatrick B, Jackson C, Kerr SM, et al. Cohort Profile: Generation Scotland: Scottish Family Health Study (GS:SFHS). The study, its participants and their potential for genetic research on health and illness. International journal of epidemiology. 2013;42(3):689–700.

14. Pidsley R, Cc YW, Volta M, Lunnon K, Mill J, Schalkwyk LC. A data-driven approach to preprocessing Illumina 450K methylation array data. BMC genomics. 2013;14:293.

15. Collaborators GDaI. Global burden of 369 diseases and injuries in 204 countries and territories, 1990-2019: a systematic analysis for the Global Burden of Disease Study 2019. Lancet (London, England). 2020;396(10258):1204–22.

16. Organization WH. Global Health Estimates 2020: deaths by cause, age, sex, by country and by region, 2000–2019. Geneva: World Health Organization. 2020.

17. Collaborators GA. Global, regional, and national burden of diseases and injuries for adults 70 years and older: systematic analysis for the Global Burden of Disease 2019 Study. BMJ (Clinical research ed). 2022;376:e068208.

18. Moorman J, Akinbami L, Bailey C, Zahran H, King M, Johnson C. Vital & health statistics. Series 3, Analytical and epidemiological studies. 35. US Dept. of Health and Human Services. Public Health Service, National Center for Health Statistics. 2012:2001–10.

19. Levey AS, Stevens LA, Schmid CH, Zhang YL, Castro AF, 3rd, Feldman HI, et al. A new equation to estimate glomerular filtration rate. Annals of internal medicine. 2009;150(9):604–12.

20. Zhang F, Chen W, Zhu Z, Zhang Q, Nabais MF, Qi T, et al. OSCA: a tool for omic-data-based complex trait analysis. Genome Biology. 2019;20(1):107.

21. Saffari A, Silver MJ, Zavattari P, Moi L, Columbano A, Meaburn EL, et al. Estimation of a significance threshold for epigenome-wide association studies. Genetic epidemiology. 2018;42(1):20–33.

22. Houseman EA, Accomando WP, Koestler DC, Christensen BC, Marsit CJ, Nelson HH, et al. DNA methylation arrays as surrogate measures of cell mixture distribution. BMC Bioinformatics. 2012;13(1):86.

23. Bollepalli S, Korhonen T, Kaprio J, Anders S, Ollikainen M. EpiSmokEr: a robust classifier to determine smoking status from DNA methylation data. Epigenomics. 2019;11(13):1469–86.

24. Zhang H, Ahearn TU, Lecarpentier J, Barnes D, Beesley J, Qi G, et al. Genome-wide association study identifies 32 novel breast cancer susceptibility loci from overall and subtype-specific analyses. Nat Genet. 2020;52(6):572–81.

25. Tcheandjieu C, Zhu X, Hilliard AT, Clarke SL, Napolioni V, Ma S, et al. Large-scale genome-wide association study of coronary artery disease in genetically diverse populations. Nature Medicine. 2022;28(8):1679–92.

26. Sakornsakolpat P, Prokopenko D, Lamontagne M, Reeve NF, Guyatt AL, Jackson VE, et al. Genetic landscape of chronic obstructive pulmonary disease identifies heterogeneous cell-type and phenotype associations. Nature Genetics. 2019;51(3):494–505.

27. Wuttke M, Li Y, Li M, Sieber KB, Feitosa MF, Gorski M, et al. A catalog of genetic loci associated with kidney function from analyses of a million individuals. Nature Genetics. 2019;51(6):957–72.

28. Sakaue S, Kanai M, Tanigawa Y, Karjalainen J, Kurki M, Koshiba S, et al. A cross-population atlas of genetic associations for 220 human phenotypes. Nat Genet. 2021;53(10):1415–24.

29. Jiang L, Zheng Z, Fang H, Yang J. A generalized linear mixed model association tool for biobank-scale data. Nat Genet. 2021;53(11):1616–21.

30. Min JL, Hemani G, Hannon E, Dekkers KF, Castillo-Fernandez J, Luijk R, et al. Genomic and phenotypic insights from an atlas of genetic effects on DNA methylation. Nat Genet. 2021;53(9):1311–21.

31. Giambartolomei C, Vukcevic D, Schadt EE, Franke L, Hingorani AD, Wallace C, et al. Bayesian test for colocalisation between pairs of genetic association studies using summary statistics. PLoS genetics. 2014;10(5):e1004383.

32. Battram T, Yousefi P, Crawford G, Prince C, Sheikhali Babaei M, Sharp G, et al. The EWAS Catalog: a database of epigenome-wide association studies [version 2; peer review: 2 approved]. Wellcome Open Research. 2022;7(41).

33. Al Rajabi A, Booth SL, Peterson JW, Choi SW, Suttie JW, Shea MK, et al. Deuterium-labeled phylloquinone has tissue-specific conversion to menaquinone-4 among Fischer 344 male rats. The Journal of nutrition. 2012;142(5):841–5.

34. Welsh J, Bak MJ, Narvaez CJ. New insights into vitamin K biology with relevance to cancer. Trends in molecular medicine. 2022;28(10):864–81.

35. Yan L, Li Q, Sun K, Jiang F. MiR-4644 is upregulated in plasma exosomes of bladder cancer patients and promotes bladder cancer progression by targeting UBIAD1. American Journal of Translational Research. 2020;12(10):6277.

36. Terkelsen T, Russo F, Gromov P, Haakensen VD, Brunak S, Gromova I, et al. Secreted breast tumor interstitial fluid microRNAs and their target genes are associated with triple-negative breast cancer, tumor grade, and immune infiltration. Breast Cancer Research. 2020;22(1):73.

37. Akter S, Choi TG, Nguyen MN, Matondo A, Kim JH, Jo YH, et al. Prognostic value of a 92-probe signature in breast cancer. Oncotarget. 2015;6(17):15662–80.

38. Ianza A, Sirico M, Bernocchi O, Generali D. Role of the IGF-1 axis in overcoming resistance in breast cancer. Frontiers in Cell and Developmental Biology. 2021;9:641449.

39. Murphy N, Knuppel A, Papadimitriou N, Martin RM, Tsilidis KK, Smith-Byrne K, et al. Insulin-like growth factor-1, insulin-like growth factor-binding protein-3, and breast cancer risk: observational and Mendelian randomization analyses with ∼430 000 women. Annals of oncology : official journal of the European Society for Medical Oncology. 2020;31(5):641–9.

40. Wright KD, Miller BS, El-Meanawy S, Tsaih S-W, Banerjee A, Geurts AM, et al. The p52 isoform of SHC1 is a key driver of breast cancer initiation. Breast Cancer Research. 2019;21(1):74.

41. Gadd DA, Hillary RF, McCartney DL, Zaghlool SB, Stevenson AJ, Cheng Y, et al. Epigenetic scores for the circulating proteome as tools for disease prediction. Elife. 2022;11.

42. Kuźnar-Kamińska B, Mikuła-Pietrasik J, Mały E, Makowska N, Malec M, Tykarski A, et al. Serum from patients with chronic obstructive pulmonary disease promotes proangiogenic behavior of the vascular endothelium. European review for medical and pharmacological sciences. 2018;22(21):7470–81.

43. Kuznar-Kaminska B, Mikula-Pietrasik J, Ksiazek K, Batura-Gabryel H. Chemokines CXCL12 and CCL21 may contribute to the development of lung cancer in COPD patients. European Respiratory Journal. 2013;42(Suppl 57):P553.

