## Additional file 1 - Supplementary Methods for "Blood-based epigenome-wide analyses on the prevalence and incidence of nineteen common disease states"

**Additional file 1 - Supplementary Methods.** The following information pertains to Supplementary Methods or Results for the manuscript '*Blood-based epigenome-wide analyses on the prevalence and incidence of nineteen common disease states'* by Hillary *et al.*

### Methylation quality control

Set 1 followed a slightly different quality control strategy to Sets 2 and 3, which together followed the same strategy. In Set 1, samples were removed if: (i) ≥1% of probes had a detection *p*-value >0.05 or (ii) there was a disagreement between self-reported sex and methylation-predicted sex. Probes were removed if: (i) ≥5% of samples had a bead count <3 or a detection *p*-value >0.05, (ii) they were non-autosomal or (iii) they overlay any SNPs and/or resided in potential cross-hybridising locations (1). In Sets 2 and 3, samples were removed if: (i) ≥0.5% of probes had a detection *p*-value >0.01 or (ii) there was a disagreement between self-reported sex and methylation-predicted sex. Probes were excluded if (i) ≥5% of samples had a bead count of 3 or less, (ii) ≥1% of samples had a detection *p*-value >0.01, (iii) they were non-autosomal or (iii) they overlay any SNPs and/or resided in potential cross-hybridising locations.

### Preparation of phenotypes

Where possible, disease states were included in both prevalent (cross-sectional) and incident (longitudinal) analyses. However, not all of the 19 disease states were present on self-report questionnaires at GS baseline. We used self-report data on twelve of the 19 disease states for our cross-sectional analyses. The remaining seven conditions that lacked appropriate self-report data at baseline were Alzheimer's dementia (AD), chronic kidney disease (CKD), inflammatory bowel disease, liver cirrhosis, ovarian cancer and both COVID-19 phenotypes (i.e. severity and long COVID). We instead used self-reported parental history of AD as a proxy variable for prevalent AD owing to the age profile of our cohort (i.e. predominantly mid-life) which is justified by the near-unit genetic correlation between family history of AD and late-onset AD (2). Furthermore, we estimated glomerular filtration rate (eGFR) from serum creatinine levels using the CKD-EPI equation and inferred CKD prevalence from the resultant data (3). Individuals with an eGFR <60 ml/min/1.73 $m^2$ were considered to have CKD.

For the four cancer phenotypes present in our incidence analyses (breast, colorectal, lung and prostate cancer), individuals present on the Scottish Cancer Registry (SMR06) were included as cases. Individuals who were recorded on the General Acute Inpatient and Day Case - Scottish Morbidity Records (SMR01) were removed from the control set. We considered two COVID-19 outcomes: severity within those who had COVID and long COVID. COVID severity was defined as a binary outcome indicating whether the participant had been hospitalised from COVID as of October 2022. Secondary SMR01 records were used to obtain COVID-19 hospital admissions using ICD-10 codes U07.1 (lab-confirmed COVID-19 diagnosis), and U07.2 (clinically diagnosed COVID-19). Long COVID was also a binary outcome but was self-reported and obtained from a subset of GS participants who took part in the CovidLife study (4). Long COVID was defined as self-reported symptom duration >4 weeks following first infection. Whereas COVID diagnosis was ascertained via record linkage for the severity phenotype, self-reported COVID diagnosis was considered sufficient for the long COVID phenotype to ensure that consistent data sources were used to build each phenotype.

**Sensitivity analyses**

The proportional hazard assumption was tested using the cox.zph function in the *survival* package (global tests and local test for CpG sites) (5). Sensitivity EWAS were performed for significant associations to further account for family structure (i.e. relatedness). This was performed for prevalent and incident disease states, considering only those associations common to basic and fully-adjusted models. These sensitivity analyses were performed using linear mixed-effects models (for prevalent disease) or mixed-effects Cox models (for incident disease) using the lmekin and coxme functions from the R *coxme* package, respectively (version 2.2-16) (6). The same covariate strategy as the fully-adjusted stage was applied with the addition of a kinship matrix to account for relatedness.

**mQTL analyses**

The GoDMC resource examined CpG sites on the Illumina 450k array and not the EPIC array, meaning some of the CpG sites in our study lack mQTL summary statistics in GoDMC. For these sites, we used mixed linear models in fastGWA to identify mQTLs in GS (n=18,413) (7). mQTL analyses were adjusted for age, sex and batch.

**Search terms for structured literature review**

We searched MEDLINE (Ovid interface, Ovid MEDLINE in-process and other non-indexed citations and Ovid MEDLINE 1946 onwards), Embase (Ovid interface, 1980 onwards), Web of Science (core collection, Thomson Reuters) and medRxiv/bioRxiv to identify relevant articles indexed as of August 31 2022. We used the following search terms or their synonyms appropriate to each database: (“blood”.mp OR “whole blood”.mp OR “peripheral blood.mp”) AND ("EWAS" OR exp "epigenome-wide*" / OR exp “epigenome-wide association” /) AND (the disease of interest e.g. “COPD” OR “chronic obstructive pulmonary disease”). Inclusion criteria were as follows: (i) original research article, (ii) EWAS performed with blood DNAm, (iii) there were at least 20 individuals in each comparison group (i.e. cases and controls) and (iv) the study examined at least one of the 19 common disease states outlined in our study. Fifty-three unique articles met inclusion criteria.

#### Incident EWAS – associations in fully-adjusted model but not basic model

Three disease states had associations in the fully-adjusted stage that were not reflected in the restricted basic model. Liver cirrhosis was associated with cg11034763 located near *LOC100506688* on chromosome 5 ($\beta$=-0.01, $p$=9.6x$10^{-10}$). COVID severity correlated with CpG hypomethylation in five genes (*APC*, *C1orf86, LDB3*, *STOML1*, *TOP3B*, range of $p$=[8.6x$10^{-10}$, 6.1x$10^{-15}$]) and hypermethylation in two genes (*CLCF1* and *RPL27A*, $p$=5.7x$10^{-10}$ and 1.1x$10^{-13}$, respectively). The probes cg04237196 and cg14012662 near *ARL4A* and *PLEKHG3* on chromosomes 7 and 14 associated with ovarian cancer ($\beta$=0.01 and -0.01, $p$=2.1x$10^{-10}$ and 2.4x$10^{-11}$, respectively).

#### References


1. McCartney DL, Walker RM, Morris SW, McIntosh AM, Porteous DJ, Evans KL. Identification of polymorphic and off-target probe binding sites on the Illumina Infinium MethylationEPIC BeadChip. Genomics Data. 2016;9:22-4.
2. Marioni RE, Harris SE, Zhang Q, McRae AF, Hagenaars SP, Hill WD, et al. GWAS on family history of Alzheimer's disease. Translational Psychiatry. 2018;8(1):99.
3. Levey AS, Stevens LA, Schmid CH, Zhang YL, Castro AF, 3rd, Feldman HI, et al. A new equation to estimate glomerular filtration rate. Annals of internal medicine. 2009;150(9):604-12.
4. Fawns-Ritchie C, Altschul DM, Campbell A, Huggins C, Nangle C, Dawson R, et al. CovidLife: a resource to understand mental health, well-being and behaviour during the COVID-19 pandemic in the UK. Wellcome Open Research. 2021;6(176):176.
5. Therneau TM, Lumley T. Package 'survival'. R Top Doc. 2015;128(10):28-33.
6. Therneau T. coxme: Mixed effects Cox models. R package version 2.2–16. 2020. 2021.
7. Jiang L, Zheng Z, Qi T, Kemper KE, Wray NR, Visscher PM, et al. A resource-efficient tool for mixed model association analysis of large-scale data. Nature Genetics. 2019;51(12):1749-55.
