## Additional file 1 - Supplementary Figures for "Blood-based epigenome-wide analyses on the prevalence and incidence of nineteen common disease states"

**Additional file 4 - Supplementary Figures.** The following information pertains to Supplementary Figures for the manuscript '*Blood-based epigenome-wide analyses on the prevalence and incidence of nineteen common disease states*' by Hillary *et al.*

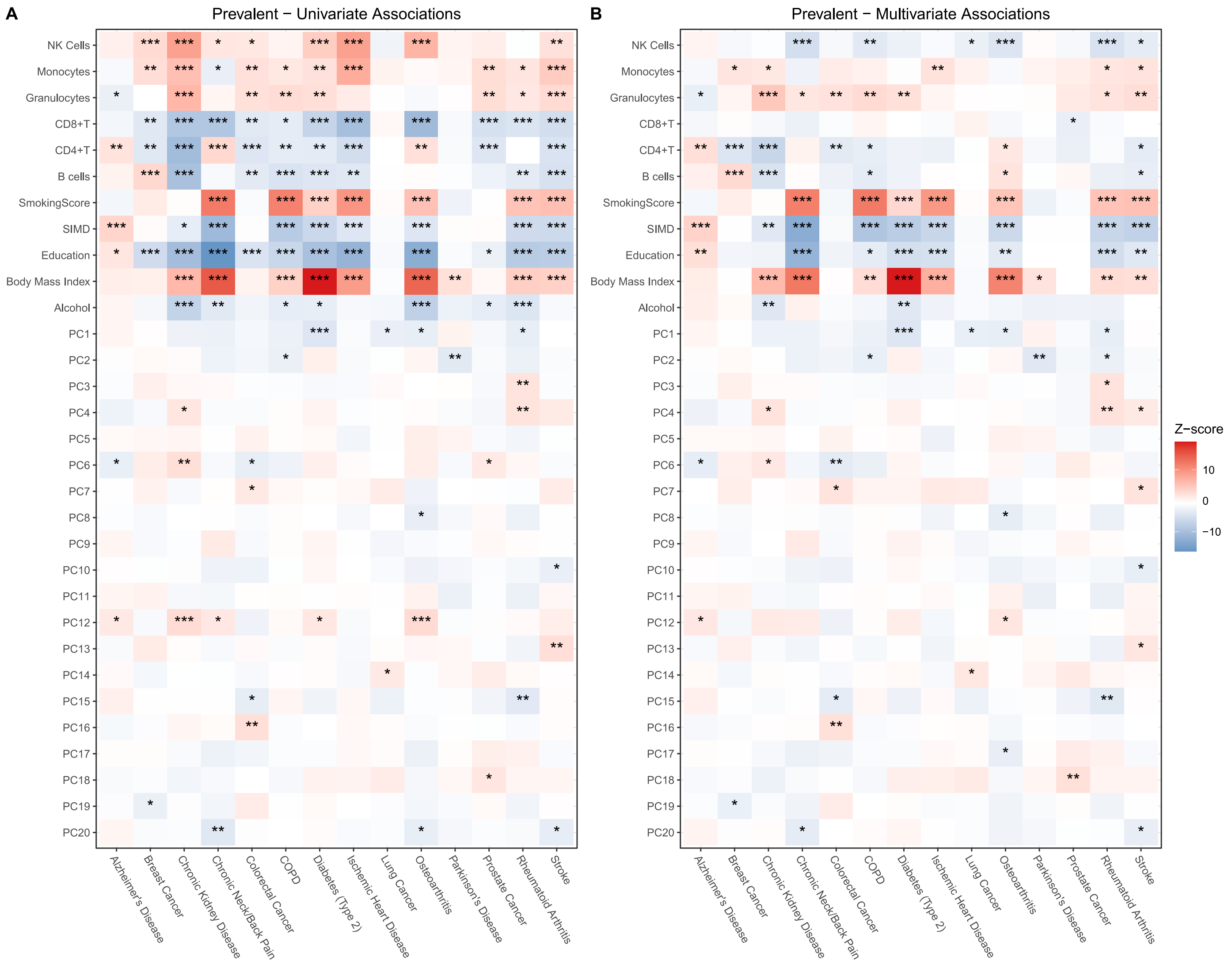


**Fig S1. Associations between covariates and prevalent disease states in univariate and multivariate logistic regression models.** Univariate models considered only the covariate and disease state as a binary outcome. Multivariate models were additionally adjusted for age and sex to obtain regression coefficients. These data are graphical representations of the data shown in **Additional file 3 – Table S4**. Stars denote the following levels of significance: *, $p<0.05$; **, $p<0.01$; ***, $p<0.001$. SIMD, Scottish Index of Multiple Deprivation.

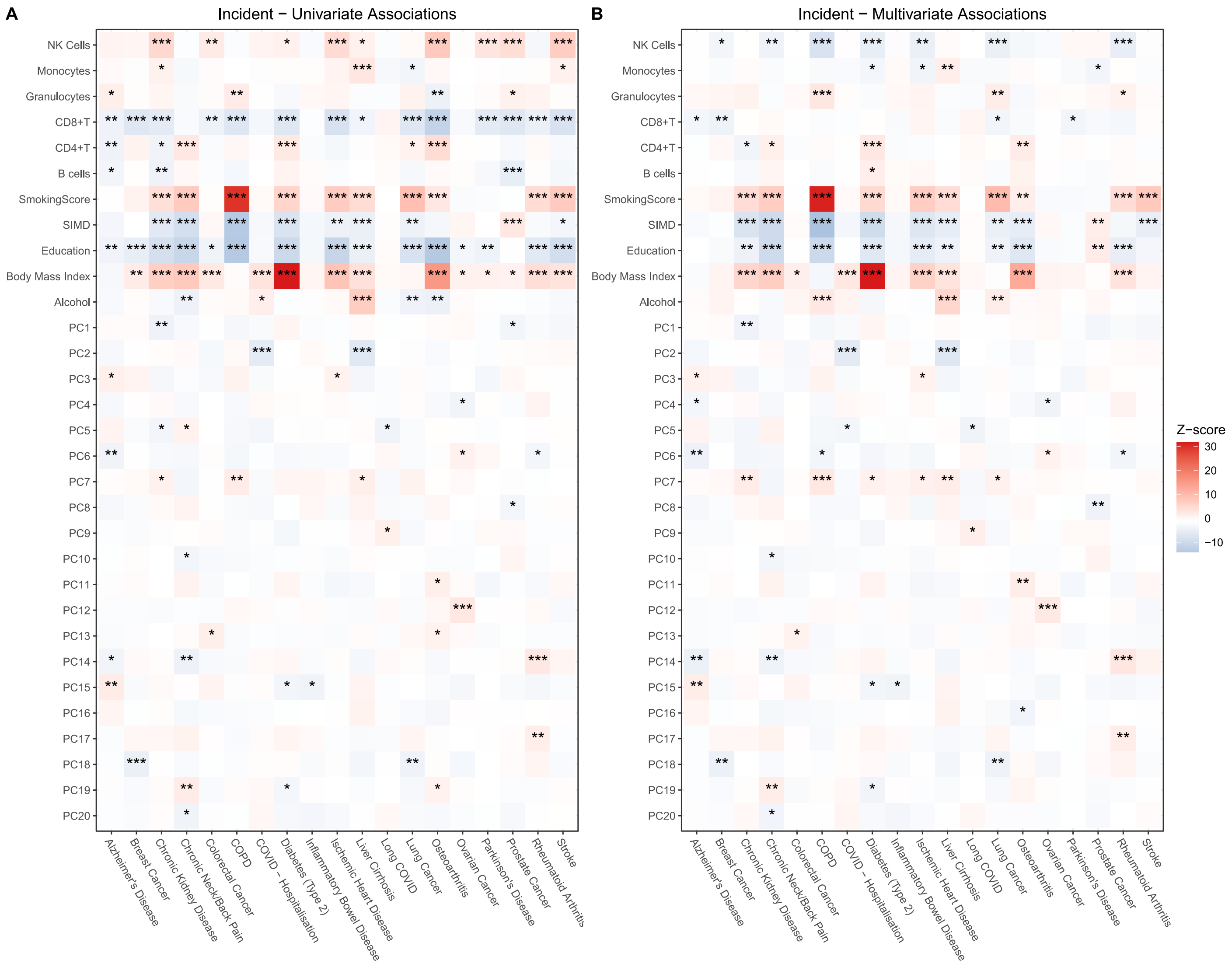


**Fig S2. Associations between covariates and incident disease states in univariate and multivariate Cox proportional-hazards models.** Univariate models considered only the covariate and disease state. Multivariate models were additionally adjusted for age and sex to obtain regression coefficients. These data are graphical representations of the data shown in **Additional file 3 – Table S5**. Stars denote the following levels of significance: *, *p*<0.05; **, *p*<0.01; ***, *p*<0.001. SIMD, Scottish Index of Multiple Deprivation.

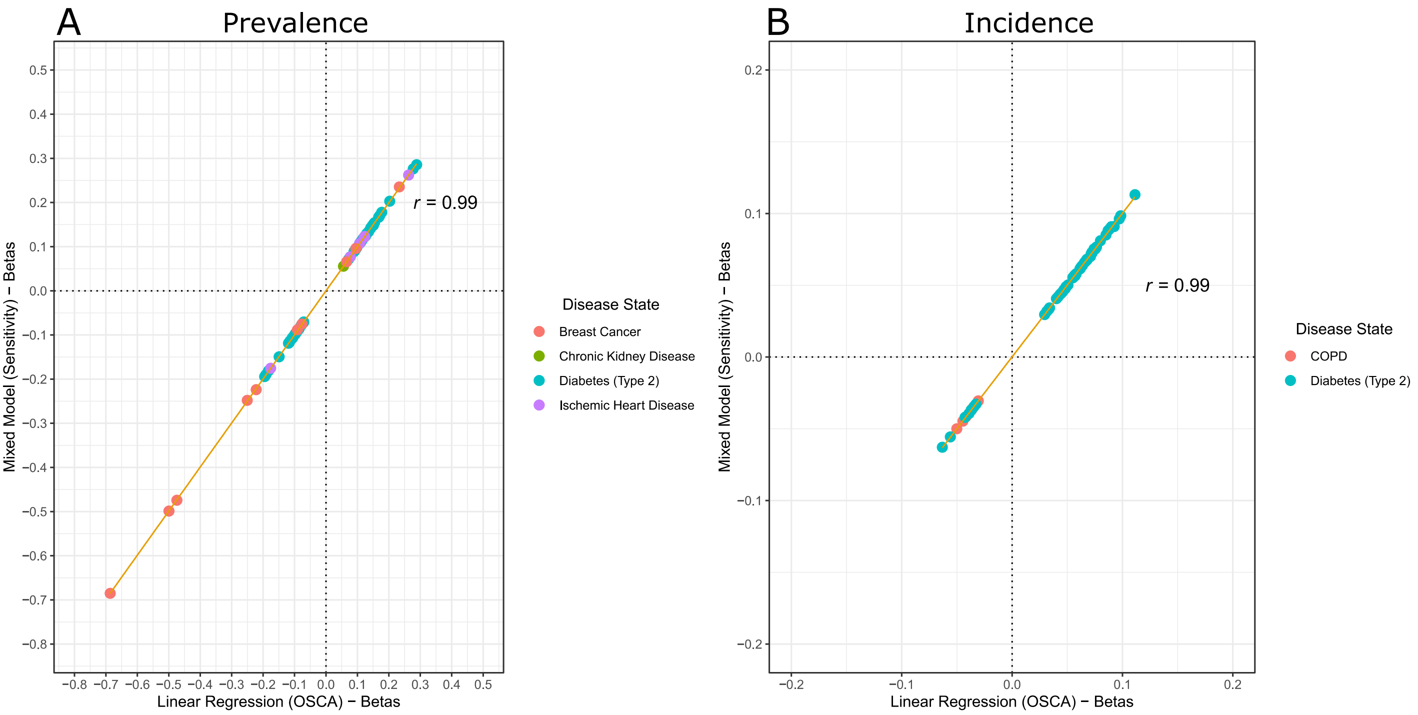


**Fig S3. Correlation between effect sizes from linear regression EWAS and sensitivity linear mixed effects analyses that further accounted for relatedness.** There was an almost unit correlation between effect sizes from the standard EWAS approach that included related individuals in Generation Scotland and effect sizes from mixed effects models that accounted for relatedness. Associations represent those that were common to both basic and fully-adjusted models. Disease states are annotated for prevalent (A) and incident disease (B) separately.
